# Evaluating the impact and cost-effectiveness of scaling-up HCV treatment among people who inject drugs in Ukraine

**DOI:** 10.1101/2021.12.13.21267712

**Authors:** Jack Stone, Josephine G Walker, Sandra Bivegete, Adam Trickey, Charles Chasela, Nadiya Semchuk, Yana Sazonova, Olga Varetska, Tetiana Saliuk, Frederick L Altice, Zhanna Tsenilova, Zahedul Islam, Dina Marunko, Bangyuan Wang, Ancella Voets, Revati Chawla, Peter Vickerman

## Abstract

**Introduction:** People who inject drugs (PWID) in Ukraine have a high prevalence of hepatitis C virus (HCV). Since 2015, PWID have been receiving HCV treatment, but their impact and cost-effectiveness has not been estimated.

**Methods:** We developed a dynamic model of HIV and HCV transmission among PWID in Ukraine, incorporating ongoing HCV treatment (5,933 treatments) over 2015–2021; 46.1% among current PWID. We estimated the impact of these treatments and different treatment scenarios over 2021-2030: continuing recent treatment rates (2,394 PWID/year) with 42.5/100% among current PWID, or treating 5,000/10,000 current PWID/year. We also estimated the treatment rate required to decrease HCV incidence by 80% if preventative interventions are scaled-up or not. Required costs were collated from previous studies in Ukraine. We estimated the incremental cost-effectiveness ratio (ICER) of the HCV treatments undertaken in 2020 (1,059) by projecting the incremental costs and disability adjusted life years (DALYs) averted over 2020-2070 (3% discount rate) compared to a counterfactual scenario without treatment from 2020 onwards.

**Results:** On average, 0.4% of infections among PWID were treated annually over 2015-2021, without which HCV incidence would have been 0.6% (95%CrI: 0.3-1.0%) higher in 2021. Continuing existing treatment rates could reduce HCV incidence by 10.2% (7.8-12.5%) or 16.4% (12.1-22.0%) by 2030 if 42.5% or 100% of treatments are given to current PWID, respectively. HCV incidence could reduce by 29.3% (20.7-44.7%) or 93.9% (54.3-99.9%) by 2030 if 5,000 or 10,000 PWID are treated annually. To reduce incidence by 80% by 2030, 19,275 (15,134-23,522) annual treatments are needed among current PWID, or 17,955 (14,052-21,954) if preventative interventions are scaled-up. The mean ICER was US$828.8/DALY averted; cost-effective at a willingness-to-pay threshold of US$3,096/DALY averted (1xGDP).

**Implications:** Existing HCV treatment is cost-effective but has had little preventative impact due to few current PWID being treated. Further treatment expansion for current PWID could significantly reduce HCV incidence.

## Introduction

Hepatitis C virus (HCV) causes substantial global morbidity[1]. The prevalence of HCV is high in people who inject drugs (PWID)[2], especially in Eastern Europe which also has the highest regional prevalence of injecting drug use (1.3% of adults[2]). The introduction of highly curative direct acting antiviral (DAA) medications for HCV[3, 4] prompted the World Health Organisation (WHO) to develop a Global Health Strategy to eliminate HCV, setting specific treatment and prevention targets to reduce HCV incidence by 80% and HCV-related mortality by 65% by 2030.

In Ukraine, national surveys of PWID estimate that the HCV prevalence among PWID is high (64% in 2017[5]), with HCV incidence also being high[6]. This high incidence and prevalence has persisted despite scale-up of harm reduction interventions[7], although coverage remains below global recommendations[8].

Ukraine initiated a National Hepatitis Program in 2019 which aims to treat 90% of those infected with HCV by 2030[9]. For this program to tackle the ongoing HCV epidemic in Ukraine, it needs to target PWID because injecting drug use (IDU) is the main driver of HCV transmission in Ukraine[10]. Numerous waves of HCV treatment have been undertaken in Ukraine since 2015, mainly supported through international donors and amongst HIV-HCV co-infected individuals, with 5,933 among current or former PWID[11–13].

We developed a mathematical model of HIV and HCV transmission among PWID to determine:

1. The impact of existing or scaled-up levels of HCV treatment.
2. The levels of treatment needed to achieve HCV elimination targets.
3. The cost-effectiveness of HCV treatments initiated in 2020.

## Methods

### Model Description

We adapted an existing dynamic, deterministic model of HIV and HCV transmission among PWID[14] to include HCV treatment. The model incorporates injecting transmission of HIV and HCV, sexual transmission of HIV, and tracks individuals following injecting cessation (ex-PWID) to fully capture HIV/HCV related morbidity. Model schematics are in Figure 1 and Supplementary Figure 1.

**Figure 1a:**
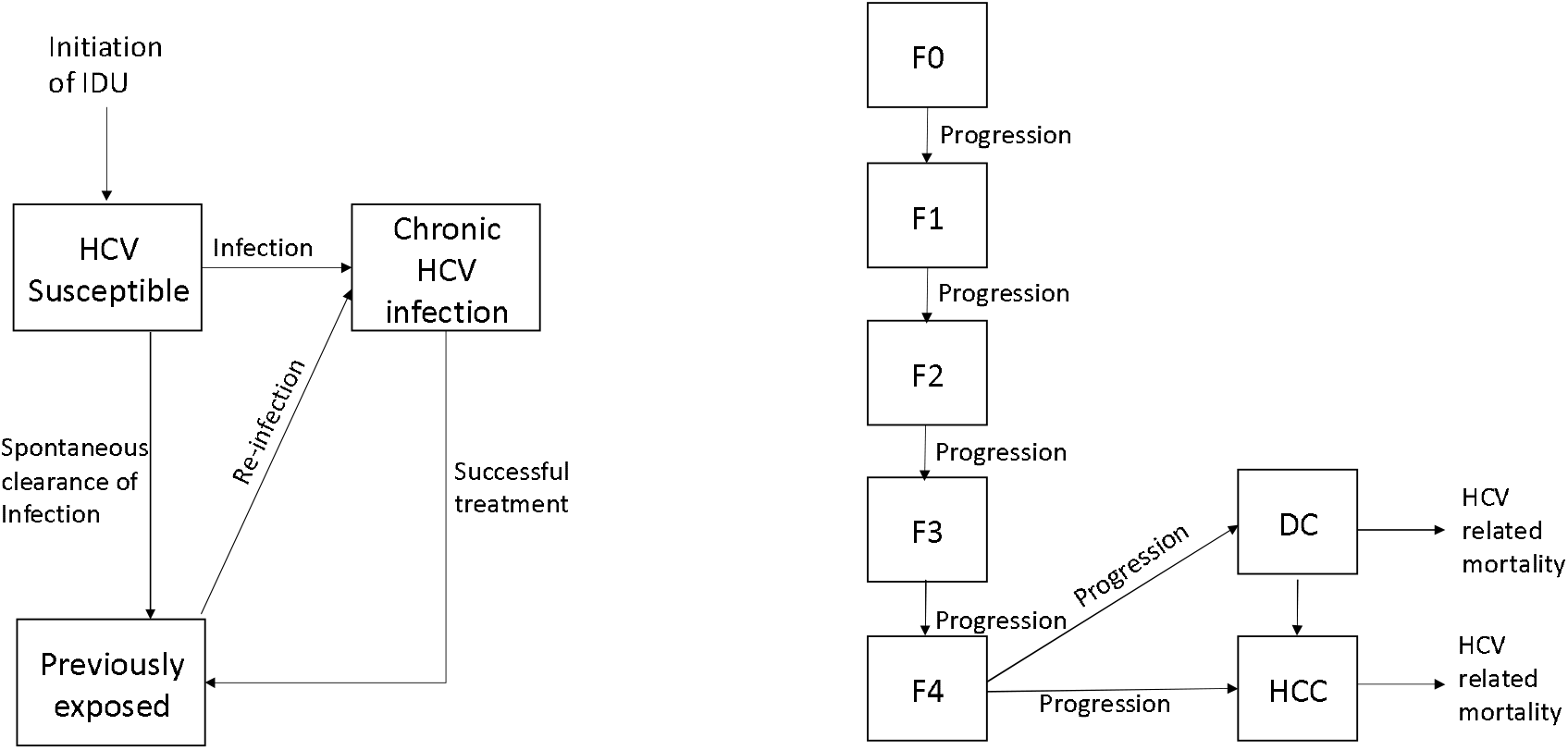
Model schematic of HCV transmission, treatment, and disease progression. DC = decompensated cirrhosis. HCC = hepatocellular carcinoma.

**Figure 1b:**
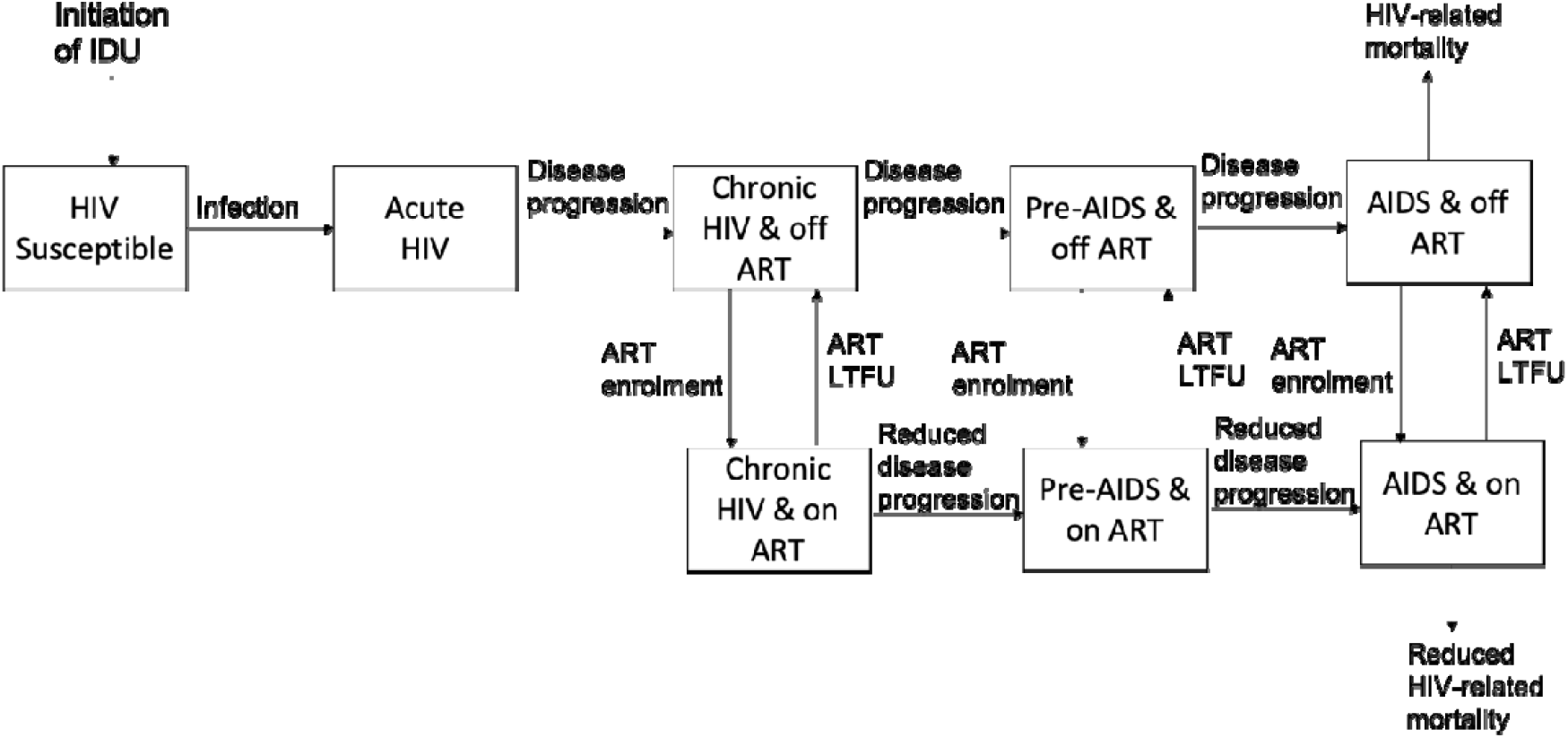
Model schematic of HIV transmission, treatment, and disease progression. ART = antiretroviral therapy. LTFU = loss to follow-up.

Individuals continually enter the model through initiating IDU and exit the model through mortality from HIV, HCV or other background causes. The model incorporates strata for being on opioid agonist therapy (OAT) or being a client of a harm reduction intervention, with PWID entering and leaving these states. OAT affects mortality rates[15], reduces the injecting risk of HIV and HCV transmission[16, 17] and improves ART outcomes[18], while being a client of an harm reduction intervention (client of non-governmental organisation or NGO) improves rates of OAT and ART initiation and reduces the risk of HIV and HCV transmission. PWID can also be incarcerated and re-incarcerated at constant but different rates, with OAT reducing these rates[19]. Compared to other community PWID, being recently incarcerated increases the risk of HIV and HCV transmission[20], while no such condition is applied during periods of incarceration.

When a susceptible PWID becomes infected with HCV, a proportion (which is lower if HIV-infected) spontaneously clear their infection[21, 22], with the remainder developing chronic infection. A time-varying number of individuals with chronic HCV initiate treatment, with those successfully treated (SVR is achieved) becoming susceptible to infection again and the remainder remaining chronically infected. No immunity is assumed following treatment.

Individuals with chronic HCV infection progress through different stages of liver fibrosis as in figure 1c, with progression rates being elevated if they are HIV co-infected but partially reduced if on ART[23]. Individuals with compensated cirrhosis can develop decompensated cirrhosis and hepatocellular carcinoma (HCC), with individuals also developing HCC if they have decompensated cirrhosis. Individuals with decompensated cirrhosis or HCC leave the model through HCV-related mortality with the mortality rate for decompensated cirrhosis being elevated if HIV co-infected[24, 25]. Disease progression can continue at a reduced rate if cured of HCV infection if the individual has cirrhosis or beyond[26, 27].

Susceptible PWID can also be infected with HIV, with individuals progressing through various infection stages (Figure 1). Individuals can be enrolled onto ART, which extends their survival and reduces infectivity.

### Model Parameterisation and Calibration

Briefly, the model was primarily parameterised and calibrated using data from the national 2011 (n=9,069), 2013 (n=9,502), 2015 (n=9,405) and 2017 (n=10,076) Integrated Bio-Behavioural Assessment (IBBA) surveys[28–31] and the 2015 Expanding Medication-Assisted Therapy (ExMAT) bio-behavioural survey (n=1,612)[32]. The model was parameterised and calibrated without the effect of HCV treatment included as done for a previous study[14], assuming low levels of HCV treatment prior to 2017, the last data-point for calibration. Key parameters and Calibration data are summarised in Table 1.

**Table 1:** Summary of main prior parameter ranges and calibration data (most recent estimates).

| Parameter | Range | Source |
| --- | --- | --- |
| **Calibration Data** |  |  |
| PWID population size | 255,702- 474,887 | [49] |
| HIV prevalence among PWID | 22.1-22.2% | 2017 APH IBBA |
| HCV antibody prevalence among PWID | 61.6-63.9% | 2017 APH IBBA |
| Proportion of PWID in contact with NGOs | 37.6-39.3% | 2017 APH IBBA |
| Odds ratio of being in contact with NGO if HIV-positive (vs HIV negative) | 2.00-2.23 | 2013/15/17 APH IBBA |
| Odds ratio of being in contact with NGO if <25 years old (vs ≥25 years old) | 0.42-0.48 | 2013/15/17 APH IBBA |
| Proportion of PWID currently on OAT | 4.4-5.3% | 2017 APH IBBA |
| Odds ratio of being on OAT if in contact with NGOs | 6.75-9.47 | 2015/17 APH IBBA |
| Proportion of HIV positive PWID on ART | 35.3-47.2% | 2017 APH IBBA |
| Odds ratio of being on ART if in contact with NGOs (vs not in contact) | 2.72-3.39 | 2015/17 APH IBBA |
| **Parameters** |  |  |
| Average duration of injecting (years) | 7.5 – 50 | 2011/13/15/17 APH IBBA's |
| Non-disease related death rate among PWID (per 100py) | 1.99 - 7.14 | [50] |
| Average length of each incarceration episode (months) | 13 – 15 | 2011/13/15/17 APH IBBA's; EXMAT |
| Rate of loss to care from ART (per 100 py) | 10.9– 15.8 | CPH HIV treatment database |
| Proportion of PWID on ART who are virally suppressed | 49-77% | [51] |
| Relative injecting risk if an NGO client vs not | 0.55-0.97 | 2011/13/15/17 APH IBBA's |
| OR of using a condom if NGO client vs not | 1.24 – 1.43 | 2011/13/15/17 APH IBBA's |
| Rate of loss to care from OAT if on OAT for <2 years (per year) | 0.45 – 0.50 | Estimated using data from [52] |
| Rate of loss to care from OAT if on OAT for ≥2 years (per year) | 0.1 – 0.15 |  |
| Relative risk of starting ART if on OAT vs not on OAT | 1.50 – 2.33 | [18] |
| Odds ratio of being virally suppressed among those on ART if on OAT vs not on OAT | 1.21 – 1.73 | [18] |
| Relative risk of ART loss to care if on OAT vs not on OAT | 0.63 – 0.95 | [18] |
| Relative risk of HCV transmission through IDU if on OAT vs not on OAT | 0.40 – 0.63 | [16] |
| Relative risk of HIV transmission through IDU if on OAT vs not on OAT | 0.32 – 0.67 | [17] |
| Relative risk of incarceration if on OAT vs not on OAT | 0.58 – 0.90 | [19, 53] |
| Relative risk of non-disease related mortality if on OAT vs not on OAT | 0.28 – 0.39 | [39] |
| Relative risk of non-disease related mortality in first 4 weeks after starting OAT vs rest of time on OAT | 0.94 – 4.10 | [15] |
| Relative risk of non-disease related mortality in first 4 weeks after leaving OAT vs rest of time off OAT | 1.51 – 3.74 | [15] |

We calibrated the model using an approximate Bayesian computation sequential Monte Carlo (ABC SMC) scheme[33]. This takes an initial series of 1,000 parameter sets randomly sampled from prior distributions, and iteratively perturbs them to improve the goodness of the fit to summary statistics on the: PWID population size; proportion of PWID that are female; HIV and HCV antibody prevalences and the differences in prevalence by age, gender and incarceration status; difference in HCV antibody prevalence between HIV positive PWID and HIV negative PWID; proportion of PWID who have ever been incarcerated, or incarcerated in the last 12 months; OAT and ART coverages and differences by NGO status; the proportion of PWID that are clients of NGOs and differences by age and HIV status; and the proportion that have been clients for < 2 years. This gave rise to 1,000 model fits which were used to give the median and 95% credibility intervals (95%CrI; 2.5^th^ to 97.5^th^ percentile range) for all model projections.

#### HCV treatment Parameterisation

We modelled four waves of HCV treatment: (i) a pilot program from June 2015-December 2017 in which 1,531 PWID (current and former) were treated[11, 12]; (ii) 255 treatments of HIV-coinfected KP members from December 2017-March 2018; (iii) EQUIP study in which 755 current and former PWID were treated over March-November 2018[13, 34]; (iv) 3,392 treatments of HIV-coinfected current and former PWID over January 2020-May 2021. Treatments were assumed to occur at a constant rate over each wave with treatments assigned by age, IDU status, HIV and ART status according to Table 2. Data on the profile of patients treated was limited for waves (ii), (iii) and (iv) and so where necessary we assumed treatments were distributed similarly to the initial pilot study or in some cases randomly (operated in the model according to the relative numbers of infections in each group). For all four waves, we distribute treatments randomly by NGO status, HIV disease stage (within those HIV positive) and incarceration history, but assign specific proportions to be among OAT clients or those on ART (Table 2). We assume that the proportion of treatments resulting in SVR remained constant across the waves at 95%[11, 12, 34].

**Table 2:** HCV treatment assumptions.

|  | **Wave (i)** | **Wave (ii)** | **Wave (iii)** | **Wave (iv)** |
| --- | --- | --- | --- | --- |
| Dates | June 2015 - Dec 2017 | Dec 2017 – Mar 2018 | Mar 2018 – Nov 2018 | Jan 2020 - May 2021 |
| Number of PWID (current and ex) treated | 1,531 | 255 | 755 | 3,392 |
| Yearly equivalent treatment rate | 633.5 | 765.0 | 1006.7 | 2394.4 |
| % of treatments resulting in SVR | 95% | No data - Assume same as Wave (i) | 95.2% | 95% |
| **Profile of those treated** |  |  |  |  |
| Number of current PWID (excluding those on OAT) | 422 (30.9% of those not on OAT) | Assume same % as Wave (i): 73.5 | Assume same % as Wave (i): 99.2 | Assume same % as Wave (i): 861.8 |
| Number current PWID on OAT | 165 | 17 | 434 | 603 |
| Total Number of current PWID | 587 (38.3% of all PWID treated) | 90.5 (35.5% of all PWID treated) | 533.2 (70.6% of all PWID treated) | 1564.8 (46.1% of all PWID treated) |
| % Male* | 73.1% | Distribute randomly | 66.0% | Distribute randomly |
| % >25 years old* | 100% (average age of 40) | Assume same as Wave (i) | 100% (IQR of those treated was 35-46) | Assume same as Wave (i) |
| % HIV positive* | 73.4% | 100% | 55.5% | 100% |
| % On ART among HIV positives* | 97.1% | Assume 100%; most HIV positives will be treated in ART clinics | 100% | 98.9% |
| HCV disease distribution* | F1: 5.23%;<br>F2: 46.9%;<br>F3: 26.1%;<br>F4: 21.8% | Distribute randomly | F1: 35.9%;<br>F2: 46.1%;<br>F3: 9.8%;<br>F4: 8.2% | Distribute randomly |
| % of treatments in prison* | 0 | 19.6% (50/255) | 0 | 10.2% (347/3392) |

\* of all PWID treated (current and ex-PWID)

#### Costs and Health Utilities

We used previously collated unit costs for the annual costs of OAT, NGO access (including NSP), ART, HIV case management, and one-time costs of HIV psychosocial services[14]. HCV treatment costs were estimated based on the 2018 study by EQUIP[13], which evaluated the cost of treating the first 522 patients enrolled in wave (iii) with sofosbuvir/ledipasvir (SOF/LDV) for 12 weeks, with weight-based ribavirin added for treatment experienced cirrhotic genotype 1 and 4 patients and all genotype 3. The intervention included 6 counselling visits with a social worker as part of a Community Based Treatment model of care. Of the patients, 85% were PWID, 52% were co-infected with HIV, and 9% had cirrhosis. RT-PCR confirmatory testing and genotype testing was costed based on the test performed at the Synevo central laboratory in Kyiv. As in EQUIP’s study, in the base case, we assumed generic pricing of SOF/LDV at $1.06 USD/pill ($89 for a 12-week treatment regimen). We excluded costs related to HIV monitoring and treatment which were included in the original EQUIP study as ART costs were estimated separately. The costs presented here are the average treatment costs across all 522 patients. The unit cost estimates and their sources are shown in Table 3.

**Table 3:** Unit costs (in 2018 USD) and disability weights used in cost-effectiveness analyses.

| Value | Value with uncertainty range | Source/Comments |
| --- | --- | --- |
| **Unit Costs (per patient)** |  |  |
| Cost of screening HCV negative patients to identify one HCV positive patient | \$1.37 per antibody test; \$33.71 per PCR test (range 27.54 to 69.61 which is range for genexpert); | Antibody tests from APH budget. PCR costs from [13] and assumes samples are sent to an external lab for testing.<br>All individuals tested receive an antibody test, all antibody-positive receive a PCR test. The cost of PCR test for positive patients is included in HCV treatment cost. Testing cost to find one RNA positive = (Antibody test cost + Antibody prevalence * PCR test cost) / (Chronic prevalence) |
| HCV treatment cost including confirmation test | \$412 (95% CI \$409-\$415) | [13] labs 37%, drugs 25%, events 17%, overheads 21% |
| HIV negative PWID annual NGO cost | \$90 (triangular distribution \$78 to \$101, accounts for regional variation in NSP cost, condoms and HCT are constant)) | [54]Includes the cost of condoms, NSP, and HIV counselling and testing |
| HIV positive PWID annual NGO cost | \$80/year (triangular distribution \$68 to \$91, accounts for regional variation in NSP cost) | [54]Includes cost of condoms and NSP. |
| One off cost in first year of initiating ART if NGO client | \$132 (triangular distribution \$82 to \$182; range is difference between cost of case management and psychosocial services) | Cost of case management or psychosocial services for each person. Based on APH budget costs, assume this cost applies to 10% of NGO clients initiating ART as per APH budget. |
| ART annual cost | \$293.47 (triangle distribution \$280.76 to \$312.53)[55] | Estimate of \$276.50 includes drug, staff, and test costs. We added 6% overheads and uncertainty bounds based on ART costs reported elsewhere[54] |
| OAT annual cost | \$300 (triangle \$194.78 to \$379.31) | Estimate of \$300 include drugs and provision, social support and incentives given to healthcare workers to support adherence. Uncertainty bounds based on OAT costs reported elsewhere[54] |
| **Disability Weights** |  |  |
| Acute or chronic HIV infection (on/off ART) | 0.078 (triangle, 0.052 to 0.111) | [35] No weights so used weights for HIV/AIDS: receiving antiretroviral treatment |
| Pre-AIDS, off ART | 0.274 (triangle, 0.183 to 0.377) | [35] Weights for HIV: symptomatic, pre-AIDS |
| AIDS, off ART | 0.582 (triangle, 0.406 to 0.743) | [35] Weights for AIDS: not receiving antiretroviral treatment. |
| Pre-AIDS or AIDS, on ART | 0.078 (triangle, 0.052 to 0.111) | [35] Weights for HIV/AIDS: receiving antiretroviral treatment |
| Compensated cirrhosis (F4) | 0.114 (triangle, 0.078 to 0.159) | [35] No weights, so used value for moderate abdominopelvic problem. Disability weights for F0-F4 are assumed to increase linearly from 0 for F0. |
| Decompensated cirrhosis | 0.178 (triangle, 0.1213 to 0.250) | [35] Weights for decompensated liver cirrhosis. |
| Hepatocellular carcinoma | 0.451 (triangle, 0.307 to 0.600) | [35] No weights so used value for metastatic cancer. |

Disability weights were taken from the 2013 Global Burden of Disease estimates[35] (Table 3). As with previous analyses[36, 37], disability weights for other conditions had to be adapted for HCV. For coinfected individuals, disability weights were combined multiplicatively.

### Model Analyses

#### Impact projections

The calibrated model was first used to estimate the impact of existing levels of HCV treatment in Ukraine. We firstly analysed the impact of the historical HCV treatments over 2015- May 2021. We then assessed the future impact of a number of treatment scenarios over May 2021-2030 with or without a concurrent scale-up in OAT (to 20% or 40% coverage) and NGOs (to 60%) among PWID in the community. The future scenarios considered were: continue the recent rate of HCV treatment (2,394/year) with either 42.5% (scenario S1) or 100% (S2) of treatments among current PWID, independent of HIV status; approximately double (S3) or quadruple (S4) the rate of HCV treatment to 5,000 or 10,000 per year, with all treatments among current PWID, independent of HIV status. For each scenario, impact was measured in terms of relative decreases in HCV incidence and chronic prevalence compared to a counterfactual in which no HCV treatment occurs over 2016-May 2021 for historic projections or over May 2021-2030 for future projections. Lastly, we estimated the annual treatment rates required to achieve the WHO target of an 80% reduction in incidence by 2030 if OAT and NGO coverage remained stable or scaled up in the community.

#### Cost-effectiveness analysis

We estimated the cost-effectiveness of the most recent wave of HCV treatment (wave (iv) - Jan 2020 – May 2021) by comparing a scenario in which all historical HCV treatments occurred up to May 2021 (‘HCV treatment scenario’), but stopped after that, with a counterfactual scenario where no HCV treatment occurred from the start of 2020 onwards (‘No HCV treatment scenario’). Costs and utilities were discounted at an annual rate of 3%. The incremental cost-effectiveness ratio (ICER) was estimated in terms of the discounted incremental costs divided by the discounted incremental DALYs averted, all measured over 2020-2070 (Time horizon 50 years). The ICER was compared to the national GDP for Ukraine (US$3,096) and to a threshold of 50% of GDP (US$1,548) which is the lowest estimated WTP threshold for Ukraine based on health opportunity costs[38]. Cost-effectiveness acceptability curves were plotted to determine the proportion of simulations that are cost-effective as a function of WTP threshold.

We performed multiple univariate sensitivity analyses to test the impact of assumptions on the ICER. These included: incorporating a cost of HCV disease-related care (Assuming 0.14% with pre-cirrhosis, 0.69% with compensated cirrhosis, and 40% with decompensated cirrhosis access care, at costs of $223, $316, and $631, respectively, based on Georgian uptake and costs); reducing the cost of DAA drugs to 50% of baseline; excluding the cost of HCV genotyping and reducing PCR test costs by 50%; calculating DALYs by taking the maximum of the HIV and HCV disability weights (rather than compounding multiplicatively); changing the time horizon to 30 years (Baseline: 50 years); changing the discount rate to 0% or 5% p.a. (Baseline: 3% p.a.); changing the SVR rate to 85% (Baseline: 95%); changing the proportion of treatments provided to current PWID to 100% (Baseline: 42.5%), with all treatments given to HIV co-infected PWID or in another sensitivity analysis given randomly based on HIV status.

We also undertook an analysis of covariance (ANCOVA) across the baseline model fits to determine which parameter uncertainties contribute most to the variability in the impact, in terms of DALYs averted, and incremental costs of HCV treatment.

## Results

The calibrated models fit the HIV and HCV prevalence data well (Figure 2), with HCV prevalence projections fitting the data well in most age and gender sub-groups (Supplementary Figure 2). The model underpredicts overall prevalence because the model is calibrated to prevalence estimates by gender and age with a higher proportion of PWID assumed to be young than are captured in the IBBS.

**Figure 2:**
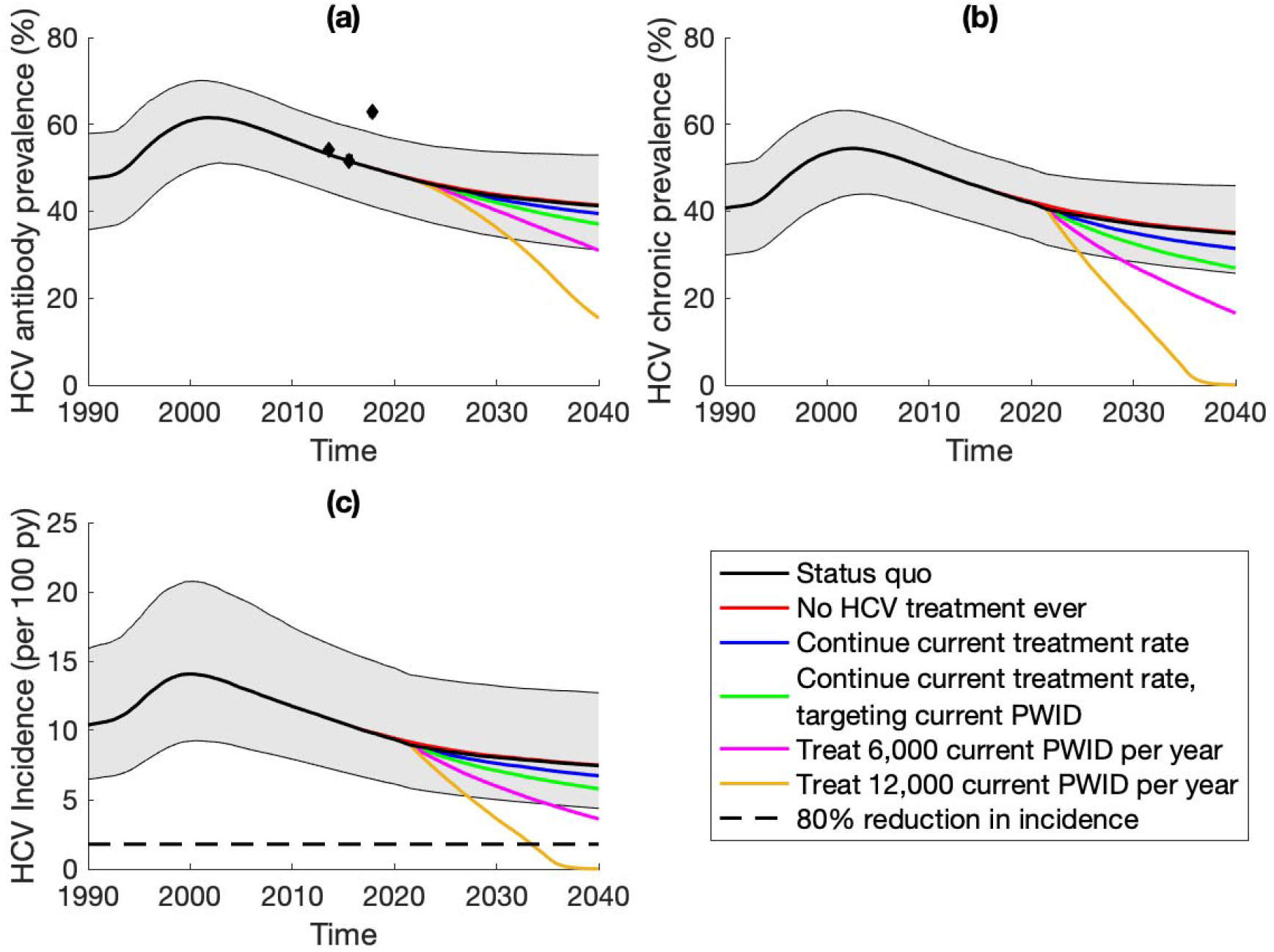
Model Projections of (a) HCV anti-body prevalence, (b) HCV chronic prevalence and (c) HCV incidence, among current PWID. Black lines and grey shaded area show the median and 95%CrI of the baseline model fits – HCV treatment up to June 2021 but no treatment after that. Coloured lines show median model projections: with no HCV treatment ever (red); continuing HCV treatment at current levels (2,394/year) with 42.5% of treatments among current PWID (blue); continuing HCV treatment at current levels (2,394/year) with all treatments among current PWID (green); scaling-up HCV treatment to 5,000/year from 2020 with all treatments among current PWID (magenta); scaling-up HCV treatment to 10,000/year from 2020 with all treatments among current PWID (yellow).

With existing levels of treatment, the model projections suggest a fairly stable HCV epidemic, with an HCV chronic prevalence of 40.5% (95%CrI: 32.2-48.6) in May 2021 and HCV incidence of 9.0 (95%CrI: 5.8-14.0) per 100 person-years (py). However, there is very little change to these epidemic trends without these treatments (figure 2), with the chronic HCV prevalence and HCV incidence among current PWID only being 2.0% (95%CrI: 1.4- 2.7) and 2.4% (95%CrI: 1.6-3.3) higher in May 2021. This is due to only 2.4% (95%CrI: 1.8- 3.1%) of chronically infected PWID being treated over 2015-2021, with most (50.7%) treatments being amongst ex-PWID.

If the treatment rates (2,394 PWID treated per year) were to continue from May 2021, then HCV chronic prevalence and incidence among current PWID would reduce by 13.5% (95%CrI: 6.8-19.5) and 15.2% (95%CrI: 7.5-21.8) by 2030 (Figure 3), respectively, if 46.1% of treatments were amongst current PWID. Conversely, if only current PWID were treated; HCV chronic prevalence and incidence would then reduce by 19.7% (95%CrI: 11.3-28.0) and 21.1% (95%CrI: 11.9-29.8) by 2030. Greater impact is achieved if OAT and NGOs are scaled-up alongside these treatments, with HCV incidence reducing by 44.9% (95%CrI: 35.0- 51.1) or 42.9% (95%CrI: 38.4-56.4) depending on whether 46.1% or 100% of treatments were among current PWID, respectively.

**Figure 3:**
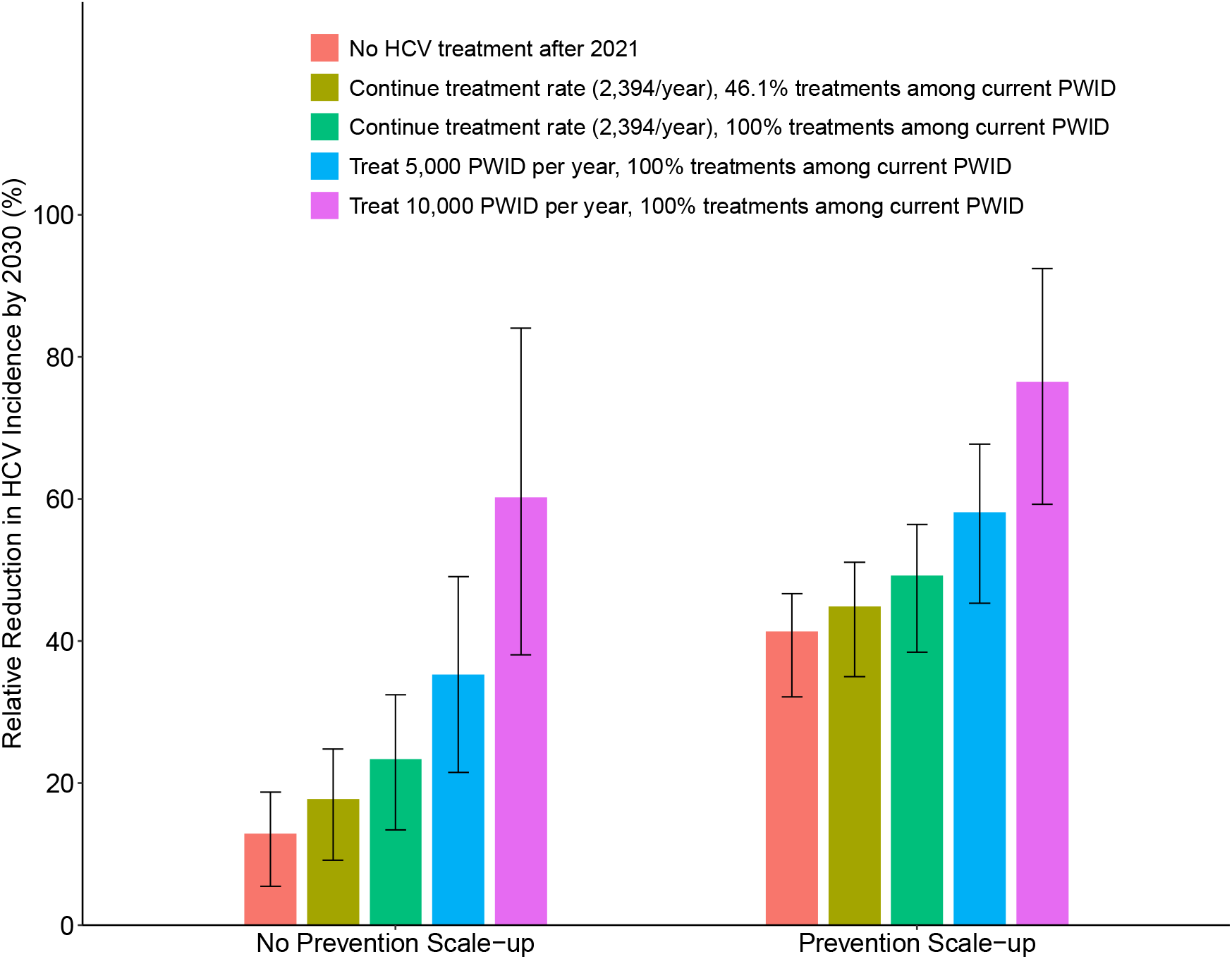
Projected impact on HCV incidence of continuing or scaling-up HCV treatment rates with or without a concurrent scale-up in prevention (OAT and NGOs scaled-up to 20% and 80%, respectively). Coloured bars show the median projection with black error bars show the 95%CrI.

**Figure 4:**
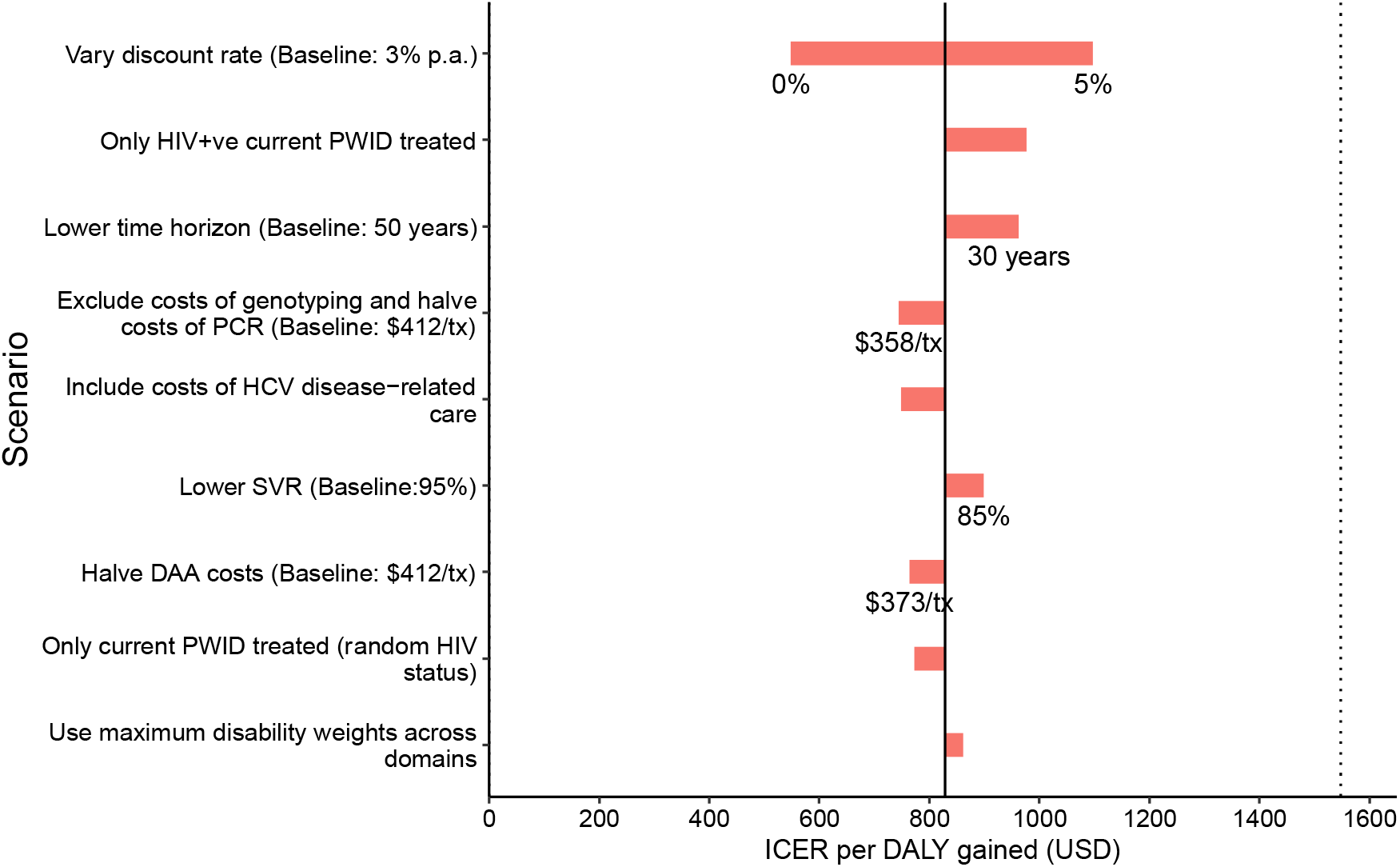
Sensitivity analyses. Red bars show the mean ICER in each of the sensitivity analyses. The solid black line shows the mean baseline ICER. The dotted black line shows the willingness to pay threshold of 0.5xGDP (US$1,548). Tx denotes treatment.

Further scaling-up treatment among current PWID could achieve greater reductions; HCV chronic prevalence and incidence would reduce by 32.5% (95%CrI: 20.0-46.1) and 33.4% (95%CrI: 20.0-47.1) by 2030 if 5,000 current PWID are treated per year or by 59.1% (95%CrI: 37.7-83.7) and 59.0% (95%CrI: 36.8-83.4) if 10,000 current PWID are treated per year. Scaling-up OAT and NGOs alongside HCV treatment could further reduce HCV incidence, with HCV incidence reducing by 58.1% (95%CrI: 45.3-67.7) or 76.5% (59.2-92.5) if 5,000 or 10,000 current PWID are treated per year, respectively (Figure 3).

To achieve an 80% reduction in HCV incidence by 2030, then 13,678 (95%CrI: 9,556- 20,365) treatments are needed per year among current PWID, decreasing to 11,045 (95%CrI: 7,674-16,959) treatments per year if NGOs and OAT are scaled-up to 60% and 20%, respectively.

### Cost-effectiveness of HCV treatment

Compared to a scenario without HCV treatment from 2020 onwards, we estimated that the status quo scenario (3,392 treatments for HIV positive PWID with 46.1% to current PWID over Jan 2020-May 2021) incurred a total incremental cost of US$2.2 million (1.86-2.59 million) over 2020-2070, including the cost of HCV treatment and additional ART costs due to HCV deaths averted. This would avert 2,651.9 (1,596.6-3,921.4) DALYs over 2020-2070, resulting in a mean ICER of US$828.8 per DALY averted (Table 4). The ICER is equivalent to 0.26 times the GDP per capita for Ukraine (US$3,096) demonstrating that this intervention is cost-effective at the 1xGDP and 0.5xGDP thresholds. In the PSA (Supplementary Figures 3 and 4), 100% of simulations were cost-effective at the 0.5xGDP thresholds. In ANCOVA, uncertainty in the HCV disease progression parameters (93.7% and 92.4% of variability in DALYs averted and incremental costs) contributed most to the variability in the DALYS averted and incremental costs. No other parameters contributed more than 5% of the variability.

**Table 4:** Cost effectiveness of HCV treatment. Table shows results using baseline cost assumptions and a discount rate of 3% per annum. 2018 USD.

|  | **No HCV treatment from**<br>**2020 onwards** | **Status Quo** | **Incremental** |
| --- | --- | --- | --- |
| **Cost of HCV treatment**<br>**(Million \$; 2019-2070)** | 0<br>(0 - 0) | 1.57<br>(1.51 - 1.59) | 1.57<br>(1.51 - 1.68) |
| **Cost of NGO**<br>**(Million \$; 2019-2070)** | 296.78<br>(228.19 - 372.38) | 296.79<br>(228.2 - 372.39) | 0.01<br>(0 - 0.03) |
| **Cost of ART**<br>**(Million \$; 2019-2070)** | 236.83<br>(142.48 - 387.73) | 237.43<br>(143.24 - 388.42) | 0.60<br>(0.31 - 0.99) |
| **Cost of OAT**<br>**(Million \$; 2019-2070)** | 111.87<br>(72.25 - 165.69) | 111.88<br>(72.27 - 165.71) | 0.01<br>(0.01 - 0.03) |
| **Total Costs**<br>**(Million \$; 2019-2070)** | 645.47<br>(499.03 - 851.71) | 647.67<br>(501.12 - 853.79) | 2.20<br>(1.86 - 2.59) |
| **DALYs** | - | - | 2,651.9<br>(1,596.6 - 3,921.4) |
| **Mean ICER**<br>**(\$ per DALY averted)** | - | - | 828.8 |

In sensitivity analyses (Figure 3), the base case ICER was most sensitive to varying the discount rate. The ICER reduced to US$547.7 per DALY averted when using a 0% discount rate but increased to US$1,097.0 per DALY averted when applying a 5% discount rate. Including potential health care savings, based on Georgian uptake and costs, removing genotyping and halving PCR test costs, or halving DDA costs had little effect (ICERs= US$743.9-764.0). In all sensitivity analyses, HCV treatment remained cost-effective at the 0.5xGDP thresholds.

## Discussion

### Main findings

Our analyses suggest that historical HCV treatments for PWID in Ukraine have had negligible impact on HCV transmission due to few treatments being given to current PWID (2.4% of current PWID were treated). Despite this, the most recent wave of treatment (Jan 2020 to May 2021) is likely to have been cost-effective ($816.4 per DALY averted), and if continued (2,394 per year) could reduce incidence by 21% by 2030 if the treatments are targeted to current PWID. HCV treatment needs to be scaled up to achieve greater impact, with the HCV elimination target of an 80% reduction in HCV incidence by 2030 being reached if 13,700 current PWID are treated each year, or 11,000 if the coverage of preventative interventions is also scaled-up.

### Strengths & limitations

The strength of our modelling includes the use of five rounds of IBBA data to parameterise and calibrate the model within a Bayesian framework that incorporates data uncertainty. The use of real-world treatment uptake, effectiveness and cost data also adds to the realism of our model projections. However, there are limitations. There was limited data on the demographics of PWID who were treated over the four waves of treatment, particularly waves (ii)-(iv). To account for this, we assumed that the same proportion of treatments were among current PWID in waves(ii)-(iv) as in wave (i), all PWID treated were >25, and that treatments were randomised across other demographics for which data was missing. Because of the relatively low number of treatments given over 2016-2021, this will not have affected our impact projections of scaling-up HCV treatment but may have affected our projections of existing impact and cost-effectiveness. Indeed, our sensitivity analysis showed that our cost-effectiveness results would improve (ICER=US$757.9) if all treatments in wave (iv) were among current PWID and they were not just targeted to HIV co-infected PWID. Also, no data was available on costs and utilisation of HCV disease related health care in Ukraine and so these cost savings from treating people were excluded in the baseline cost-effectiveness analyses making our results conservative. In a sensitivity analysis, we applied Georgian costs and utilisation data for HCV care, which only reduced the ICER by 10%, suggesting this may not be an important omission at the assumed low rates of utilisation.

### Comparisons with existing studies

Several previous modelling analyses for Ukraine have evaluated the impact and cost-effectiveness of HIV and HCV interventions for PWID; including OAT in prisons or the community[39–43], NGO programming [14, 44], or ART and PrEP[42, 43]. However, to our knowledge this is the first study to evaluate the impact and cost-effectiveness of HCV treatment among PWID in Ukraine. A recent modelling analysis for five countries in Eastern Europe and Central Asia (Belarus, Georgia, Kazakhstan, Moldova, and Tajikistan) found that increasing DAA treatment rates to 50% of those diagnosed, which ranged from 14-27% of PWID would avert between 1-15% of new HCV infections over 20 years[45] and that substantial impact would only be achieved if diagnosis rates among PWID were improved. Although the study estimated the cost-effectiveness of scaling-up DAA access and HCV diagnosis, these were only assessed in terms of life-years gained and in combination with scaling-up other interventions, finding that over 20 years a scenario with expanded OAT, NSP, ART, HCV screening, DAA access and ART could be very cost-effective or cost-saving if costs of DAAs were $900 per therapy or less. However, they did not include other costs incurred during the treatment pathway. In contrast, we used a full costing of HCV treatment in Ukraine that included costs of DAAs, RNA confirmatory testing, genotype testing, staff time, overheads, and counselling sessions, so adding realism to our cost-effectiveness projections. Although the impact projections are difficult to compare because of differences in how treatment scale-up is implemented, our results are similar in that a significant scale-up of DAA treatment is needed to substantially reduce HCV incidence.

### Implications

We have shown that HCV treatment among PWID in Ukraine is cost-effective and could have considerable impact and possibly achieve elimination if scaled up. However, with only 41% of the ∼167,000 HCV infected PWID currently aware of their infection status[31], drastic increases in HCV case-finding are needed to achieve this. In Ukraine, among those infected with HCV, PWID that have been incarcerated in the last 12 months (aOR:0.88) are less likely to be diagnosed than those who have not been recently incarcerated, whilst PWID that have been diagnosed with HIV (aOR:3.53, compared to those HIV negative or who are HIV positive but undiagnosed) or are clients of NGOs (aOR: 2.39) or have a history of OAT (aOR: 1.62 are more likely to be diagnosed than those not engaged in these interventions (unpublished analyses of IBBA data), findings which are supported by a recent international review[46]. Given the high HCV prevalence among PWID in prisons or with a history of incarceration in Ukraine, prisons may be an important setting for HCV testing in Ukraine as is the case in many global settings[47, 48]. Additionally, further efforts are needed to diagnose those who are not diagnosed with HIV, whilst scaling-up OAT and NGOs could be crucial for increasing case-finding and linking PWID to HCV treatment. Furthermore, the expansion of these cost-effective interventions could reduce the number of treatments needed for reaching HCV elimination and would provide other benefits in reducing HIV transmission and levels of mortality among PWID[14, 39].

## Data Availability

All data produced in the present study are available upon reasonable request to the authors

## Acknowledgements

This study was funded by Frontline AIDS. JS and PV also acknowledge funding from NIAID and NIDA (R01AI147490). JS, PV and FLA acknowledge funding from NIDA (R01DA033679, R21 DA047902, R01 DA029910). PV, SB and JS are also supported by the UK National Institute for Health Research (NIHR), Health Protection Research Unit (HPRU) in Behavioural Science and Evaluation at the University of Bristol.

## Notes

**Conflicts of Interest:** The Global Fund to fight AIDS, tuberculosis, and malaria (GF) or other international funders had no role in these analyses or in decisions to publish. JGW, PV and FLA report research grants from Gilead unrelated to this work and FLA has research grants from Merck unrelated to this grant.

### Competing Interest Statement

The Global Fund to fight AIDS, tuberculosis, and malaria (GF) or other international funders had no role in these analyses or in decisions to publish. JGW, PV and FLA report research grants from Gilead unrelated to this work and FLA has research grants from Merck unrelated to this grant

