## Supplementary Materials for "Evaluating the impact and cost-effectiveness of scaling-up HCV treatment among people who inject drugs in Ukraine"

Supplementary Figure 1a: Model schematic of Incarceration.

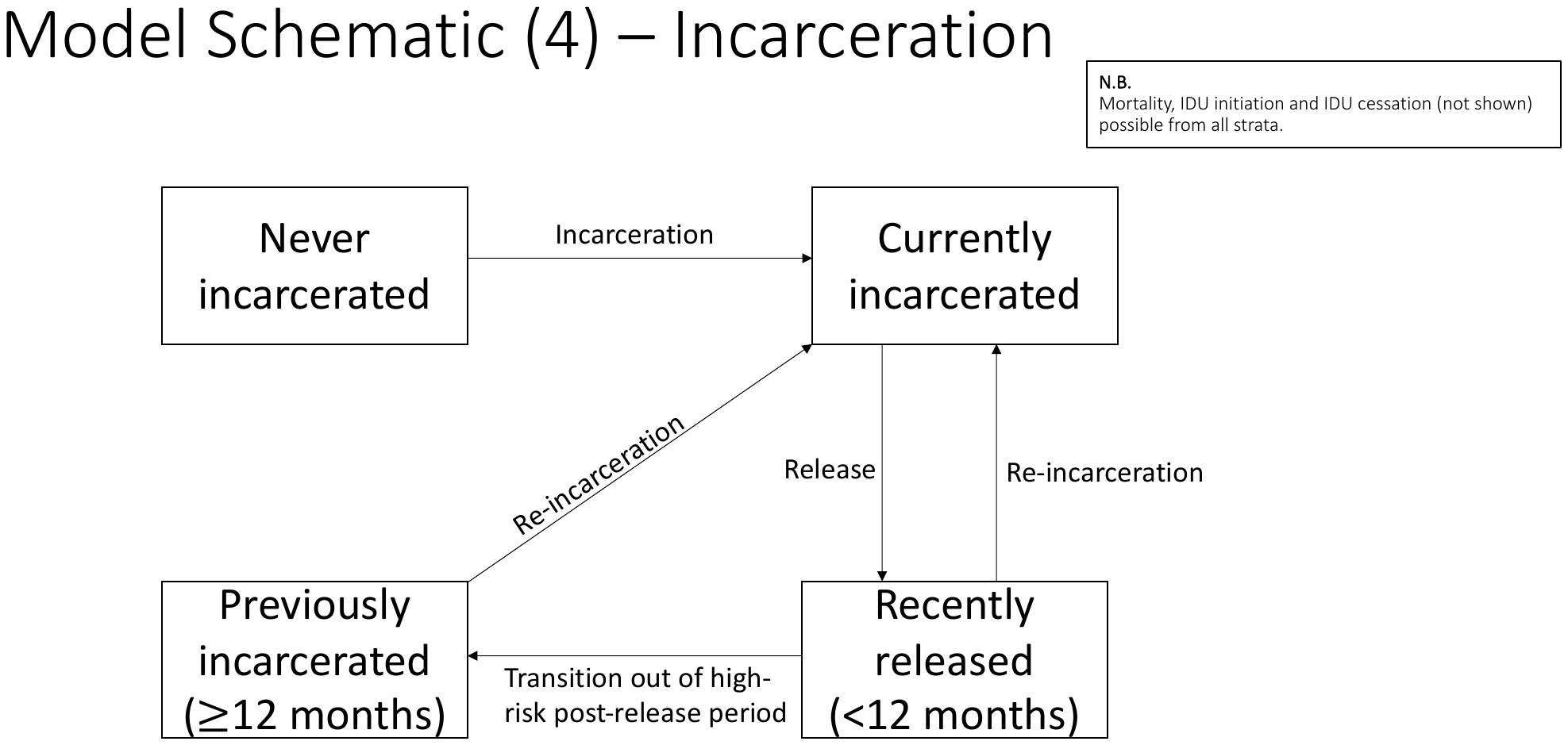

Supplementary Figure 1b: Model schematic of contact with NGO and OAT.

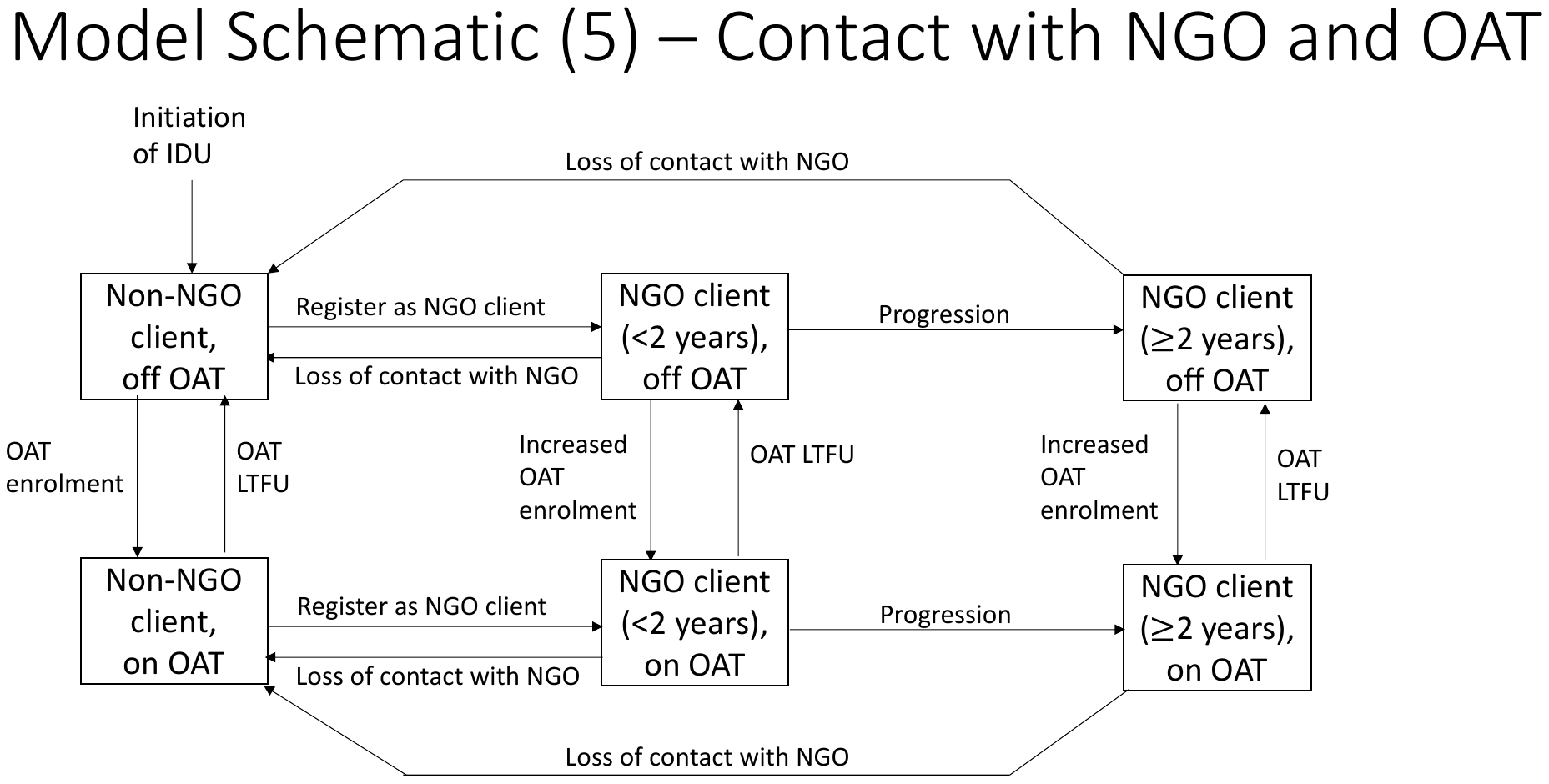

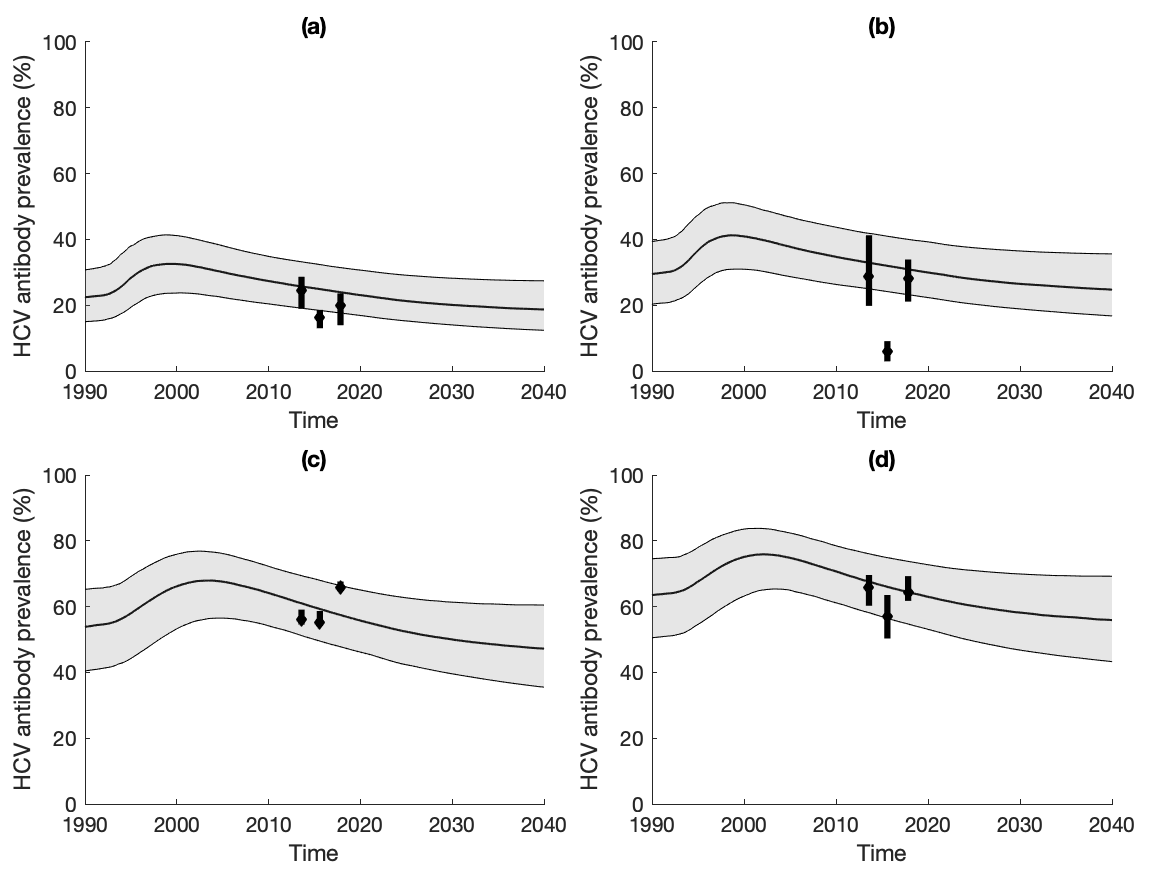

Supplementary Figure 2: *Status quo projections of community HCV antibody prevalence by age and gender. (a) young male PWID (<30); (b) young female PWID (<30); (c) older male PWID (>=30); (d) older female PWID (>=30). Black lines and grey shaded area show the median and 95%CrI of the baseline model fits. Data points with whiskers show data and their 95% CIs.*

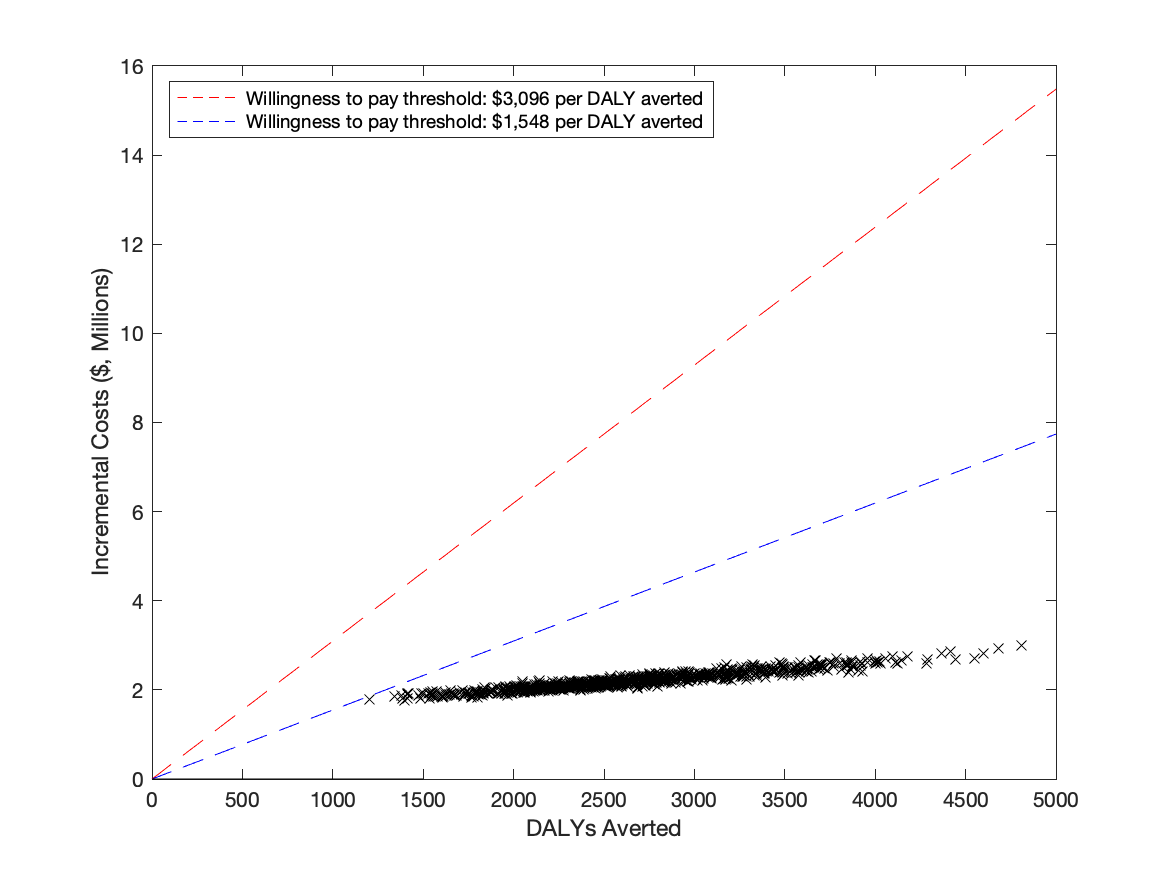

Supplementary Figure 3: *Cost-effectiveness plane for the Baseline cost-effectiveness analysis. Black points represent each model run. Dashed lines show the WTP threshold of 1xGDP (red) and 0.5xGDP (blue).*

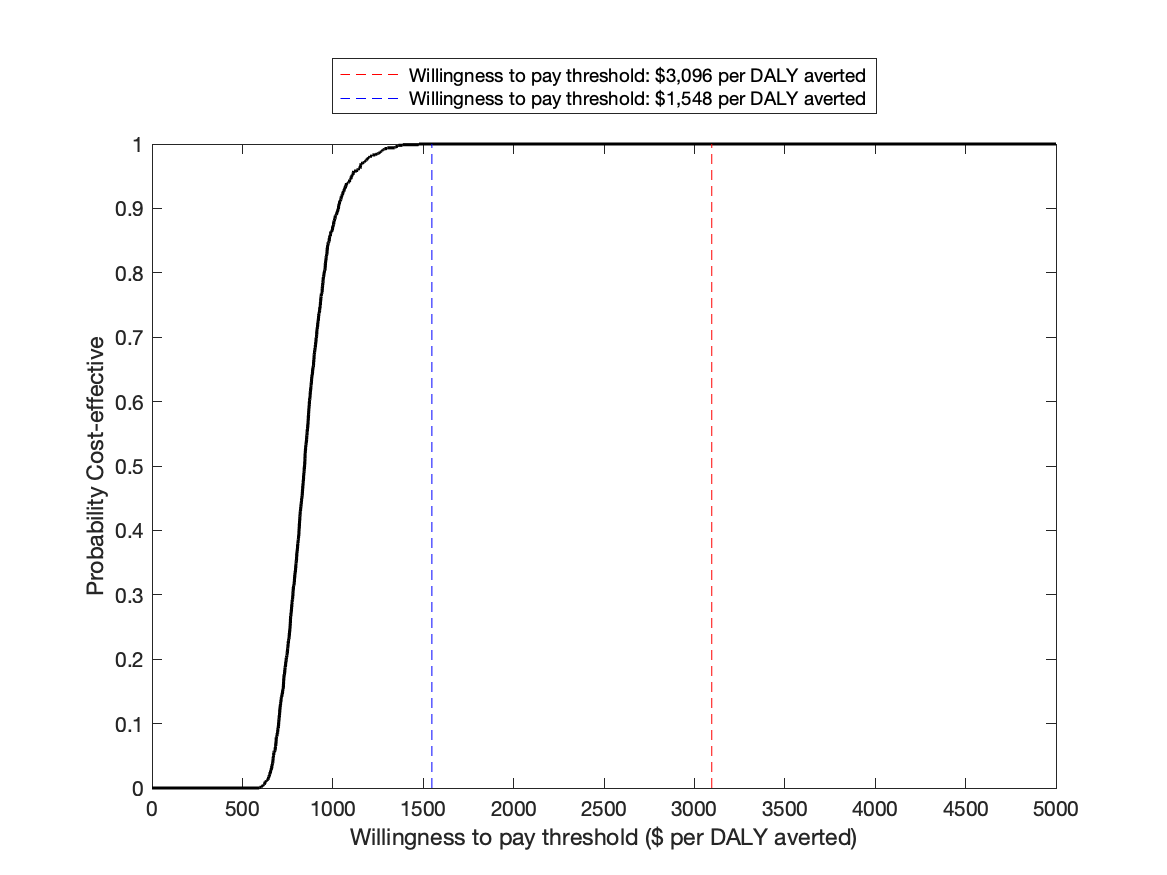

Supplementary Figure 4: *Cost-effectiveness acceptability curve for the Baseline cost-effectiveness analysis. Dashed lines show the WTP threshold of 1xGDP (red) and 0.5xGDP (blue).*
